# Application of physiological network mapping in the prediction of survival in critically ill patients with acute liver failure

**DOI:** 10.1101/2024.04.21.24306147

**Authors:** Tope Oyelade, Kevin P. Moore, Ali R. Mani

**Affiliations:** Institute for Liver and Digestive Health, Division of Medicine, UCL, London, UK; Network Physiology Laboratory, Division of Medicine, UCL, London, UK

**Keywords:** Acute Liver Failure (ALF), Network Physiology, Principal Component Analysis, Paracetamol, Acetaminophen

## Abstract

Reduced functional connectivity of physiological systems is associated with poor prognosis is critically ill patients. However, physiological network analysis is not commonly used in clinical practice and awaits quantitative evidence. Acute liver failure (ALF) is associated with multiorgan failure and mortality. Prognostication in ALF is highly important for clinical management but is currently dependent on models that do not consider the interaction between organ systems. This study is aimed to examine the impact of physiological network analysis, in prognostication of patients with ALF.

Data from 640 adult patients admitted to the ICU for paracetamol-induced ALF were extracted from the MIMIC-III database. Parenclitic network analysis was performed on the routine biomarkers and network clusters were identified using the k-clique percolation method.

Network analysis showed that the liver function biomarkers were more clustered in survivors than in non-survivors. Arterial pH was also found to cluster with serum creatinine and bicarbonate in survivors compared with non-survivors, where it clustered with respiratory nodes indicating physiologically distinctive compensatory mechanism. Deviation along the pH-bicarbonate and pH-creatinine axes could significantly predict mortality independent of current prognostic indicators. These results demonstrate that network analysis can provide pathophysiologic insight and predict survival in critically ill patients with ALF.

## Introduction

The human body is a complex system composed of an intertwined network of physiological components that interact with each other to maintain homeostasis. The study of robustness in complex physiological networks is important because it can aid in understanding disease processes and the compensatory mechanisms that lead to survival after critical illnesses such as multiple-organ failure. Our knowledge about predictors of survival in critically ill patients is limited to clinical and epidemiological studies that have identified risk factors for mortality, such as a greater number of failing organs assessed by the SOFA score (Sequential Organ Failure Assessment) [1,2]. Recent studies using various machine learning approaches have also indicated that the aggregation of previous disease history and acute physiology measures can predict in-hospital mortality in critically ill patients [3]. While all these studies are useful for prognostication in intensive care units, they rarely provide hypotheses that may lead to the development of interventions to alter the progression of the disease.

Recently, the network of physiological organ interactions has been studied within the context of critical illness [4–6]. The emerging field of network physiology has established the groundwork for understanding and quantifying global physiological behaviours arising from networked interactions across systems in health and disease [7,8]. At least in theory, mapping the physiological network during an acute pathological insult in survivors may reveal compensatory mechanisms deployed to regain homeostasis with the potential for the development of new therapies. Indeed, physiological network mapping can also improve our understanding of the pathophysiology of the disease.

The liver plays a pivotal role in physiological processes within the body, positioning it as a central hub in the control of various physiological mechanisms. Clinicians readily acknowledge the functional connectivity of the liver with other organs, which is particularly evident in patients with liver failure who manifest involvement of multiple organs, such as neural, cardiovascular, renal, and metabolic dysfunction, as well as acid/base and electrolyte imbalance [9]. Chronic liver failure (cirrhosis) is associated with impaired cardiovascular control [10–12] and thermoregulatory dynamics [13–15]. The application of a network approach in patients with cirrhosis has also revealed that organ system network disruption is associated with a poor prognosis in patients with chronic liver failure [16,17]. Specifically, there was a greater correlation between physiological biomarkers in liver cirrhosis in survivors than in non-survivors, indicating that more connected organ systems are present in survivors [16,17]. Furthermore, physiological network connectivity indices could predict 6-month survival in patients with cirrhosis independent of age and the severity of liver disease [17,18]. Such a network approach in liver cirrhosis is helpful, as it can provide prognostic information (e.g., for application in liver transplant allocation) as well as aid in predicting response to therapy (e.g., targeted albumin therapy) [18].

The process of cirrhosis is a slow process that takes more than a decade to significantly affect physiological networks. On the other hand, acute liver failure (ALF) is an acute process defined as the presence of severe worsening acute liver injury (<26 weeks) in patients with no history of chronic liver disease [19]. ALF can be caused by acute exposure to high doses of hepatotoxins such as paracetamol (acetaminophen). Paracetamol-induced ALF is the most common cause of ALF in Western countries [20]. The rapid deterioration of liver function in ALF patients is strongly associated with a high risk of mortality, which may be prevented by timely liver transplantation. However, a subpopulation of ALF patients is known to recover fully without transplantation. Due to its accidental occurrence (e.g., drug overdose), paracetamol-induced ALF often occurs in otherwise healthy individuals. These patients need to adapt and employ suitable compensatory mechanisms to survive rapidly. While ALF is a multiorgan systemic disease involving other organ systems (e.g., hepatic encephalopathy), a network approach has not yet been applied to understand the differences in organ-system interactions between survivors and non-survivors. In this study, we aimed to apply a network mapping approach to investigate the interaction between multiple variables representing various organ systems in a cohort of critically ill patients with ALF. We also aimed to compare the prognostic value of the network mapping approach with the current clinical prognostic indicators used to assess critically ill patients with ALF (e.g., the King’s College Criteria score and SOFA score).

## Methods

### Database Description and Extraction

The data analysed in this study were sourced from the third version of the Medical Information Mart for Intensive Care (MIMIC-III) following training, application, and acquisition of the required permissions. MIMIC-III data has been deidentified in accordance with Health Insurance Portability and Accountability Act (HIPAA) standards and the project approved by the Institutional Review Boards of Beth Israel Deaconess Medical Center and the MIT (IRB protocol nos. 2001P001699/14 and 0403000206, respectively). The authors involved in data extraction (TO and ARM) completed mandatory online ethics training at MIT and were credentialled (IDs 10304625 and 48067739).

The MIMIC-III dataset includes more than 53,000 unique hospital admissions. Initially, the complete MIMIC-III clinical dataset was downloaded to a secured cloud storage of the University College London (UCL), and structure query language (SQL) code was used to extract the required data based on the inclusion criteria. Thus, the inclusion criteria included being an adult (aged 18 years and above) and being diagnosed with ALF linked with paracetamol/acetaminophen overdose at the time of ICU admission. Patients with less than 50% of clinical data records or those with missing follow-up and hospital mortality records were excluded from this study.

Specifically, the SQL code extracted data of patients aged 16 years and older of any sex who were diagnosed with acute liver failure (ICD_DIAGNOSIS) based on the International Classification of Diseases 9^th^ revision (ICD9) Code (570) [21] and who had a combination of the strings ‘acetaminophen’ or ‘paracetamol’ or other known commercial names of acetaminophen-containing combination drugs as well as the string ‘overdose’ in their clinical notes (NOTEEVENTS). The combination of the clinical note details with the ICD9 code was performed to reduce the error inherently associated with the now obsolete ICD9 diagnostic code used in the MIMIC-III data, especially for acetaminophen/paracetamol-induced ALF [22]. The list of patients was then used to extract the laboratory variables (LABEVENTS) and vital variables (CHARTEVENTS). Other clinical variables of the identified patients, including age, sex, ICU length of stay, and in-hospital mortality, were also extracted for analysis.

Furthermore, the minimum Glasgow Coma Score (GCS) and King’s College Criteria (KCC), which index patients’ level of consciousness [23] and severity of ALF [24], respectively, were computed based on the available data recorded during the first day of ICU admission and included in the analysis. The GCS score was calculated based on the patient’s verbal and motor responses and eye opening [25]. For KCC, patients were scored based on acidosis (arterial pH < 7.30), coagulopathy (international normalized ratio > 6.5), kidney function (serum creatinine > 3.4 mg/dL), and the presence of hepatic encephalopathy of grade 3 or 4 according to the West Haven grading system [24]. However, because the MIMIC-III dataset does not contain the West Haven grades (or any grading system) of hepatic encephalopathy, patients whose GCS score was ≤ 8 were classified as West Haven grade III or IV according to clinical guidelines [26,27].

The sequential organ failure assessment (SOFA) score, which is often used to assess the severity (morbidity) of critically ill patients, was also calculated since the study population is primarily those admitted to the ICU [28]. Finally, mortality was defined as patients who died within 28 days of hospitalization and those who underwent liver transplantation (within 28 days) because these patients probably would not have survived if they had not undergone transplantation.

### Physiological network mapping

We applied Parenclitic network mapping for network mapping based on patients’ clinical or biochemical biomarkers. Parenclitic network mapping is a novel approach for network analysis [17] that facilitates the mapping of individual data points within models constructed from a reference population (e.g., patients who survive ALF). Initially, the correlation between clinical/biochemical biomarkers was assessed in the reference population to determine the expected relationship between the pair of biomarkers. To map the network of individual patients, a parenclitic approach was used. This analysis measures the deviations of an individual patient from the expected relationship between variables in the reference population. For a detailed exploration of Parenclitic network analysis in the context of cirrhosis, refer to the research conducted by Zhang et al. [17]. In this study, the data of patients who survived ALF 28 days post-ICU admission were used as the reference population. Regression analysis was performed on pairs of clinical variables based on a p value that was corrected for the number of comparisons (Bonferroni correction). A population network was then created based on statistically significant regressions. The parenclitic deviations from the significantly correlated models along each axis (pairs of variables) were computed and used to construct a network map of individual patients. The deviation along each axis is reported as δ-A/B (e.g., δ-chloride/bicarbonate), where A or B represents a biochemical variable (e.g., chloride and bicarbonate), and δ denotes deviation from the regression line between A and B in survivors with ALF (i.e., the reference population). Network mapping was carried out using an in-house code developed in MATLAB (MathWorks, CA, USA).

### Detection of local clusters within the network

For a complex physiological network containing a high number of nodes, understanding the cluster of tightly interconnected nodes could provide insight into the core structures within the network. The k-clique percolation method is a cluster detection technique that is robust to the overlap of shared characteristics between clusters within a network [29,30]. The technique defines all the cliques (a subgraph of a network where all member nodes are adjacent to each other) within a network that shares k-1 (at least one) nodes [31]. A clique in this instance is defined as a complete subnetwork within the overall phylogenetic network composed of physiological variables with a greater likelihood of being correlated compared with variables from other communities [32]. Thus, the physiological communities are defined as organ systems that are more closely aligned in terms of functionality compared with other nodes within the overall network. In this study, the k-clique percolation method was employed using a MATLAB function originally written by Ahn-Dung Nguyen [33]. The nodes and edges of the detected community (cliques) are color-coded for clarity and show which variables belong to the same clique.

### Principal Component Analyses

Since Parenclitic network mapping is based on the correlation between physiological variables in a reference population, it shares some statistical insight with the principal component analysis commonly used for dimension reduction. Thus, principal component analysis was also performed on patient biomarkers to assess which combination of variables (principal components, PCs) can predict survival in patients with paracetamol-induced ALF. This analytic method can identify clusters of variables that are highly correlated before dimension reduction. Therefore, it has been used for the identification of clusters during network analysis within the context of critical care [6]. Briefly, principal component analysis reduces the dimension of a dataset by computing the best combination of variables (PCs) that explains most of the variability in a dataset [34]. For this analysis, Bartlett’s test of sphericity (chi-squared) and KMO-MSA (Kaiser, Meyer, Olkin’s Measure of Sampling Adequacy) were used to assess whether the dataset was suitable for factor analysis, while Kaiser’s rule based on eigenvalues > 1.0 was used to determine relevant PCs that explained a significant portion of the variability in the data. To extract variables that could be easily interpreted, the normalized varimax rotation method was used [35].

### Statistical analysis

The characteristics of patients who survived and those who did not survive were compared with continuous and categorical variables and are presented as the mean ± standard deviation (SD) or median and interquartile range (IDR), respectively. For comparisons of continuous variables between patient groups, independent samples t tests or Mann‒Whitney U tests were used depending on whether the data were normally distributed. In general, parental deviations (δs), as well as principal components, were subjected to statistical analysis for survival predictions. The chi-squared test was used to compare categorical variables with significantly different variables (parenclitic and principal components) that were subjected to univariate and multivariate Cox regression analyses. To compare the hazard ratios of the network indices, the scales of the parenclitic deviations were normalized before Cox regression analysis using Z transformation. Receiver operating characteristic (ROC) curve analyses were performed for variables that were independently predictive of 28-day ICU mortality, and the area under the curve (AUC) was computed. Positive and negative predictive values of the ROC cut-offs were computed using the sensitivities and 1-point specificities. All the statistical analyses were performed with SPSS Statistics 26 (IBM Corp., Armonk, NY). For the assessment of prognostic improvement from the combination of parenclitic indices and principal components with the SOFA score, the Brier score, integrated discrimination improvement (IDI), and the net reclassification index (NRI) were computed with Stata statistical software (Stata/MP, Version 17.0). Essentially, lower Brier scores translate into a better predictive model, while IDI and NRI decrease misclassification due to the addition of a new variable to a predictive model [36,37]. For the interpretation of all the statistical analysis results, a two-tailed p-value less than 0.05 was used as the cut-off for significance. For the combination of the SOFA score with the independent predictive indices, composite scores were created using the formula β1 ×SOFA + β2 × δ, where β is the multivariate Cox regression coefficient of the variables.

## Results

### Patient characteristics

A total of 640 patients with ALF due to paracetamol overdose were included in this study, 249 (38.9%) of whom either did not survive a 28-day ICU stay or underwent liver transplantation during admission. Overall, there was no significant difference in age (years), sex, ICU length of stay (LOS), serum bicarbonate, serum sodium (Na), international normalized ratio (INR), white blood cell (WBC) count, heart rate (HR), respiratory rate (RespR), or arterial blood pH (pH). However, there was significantly greater alanine aminotransferase (ALT) and aspartate transaminase (AST), serum albumin, chloride, platelet count, body temperature, mean blood pressure, oxygen saturation, haemoglobin, and Glasgow Coma Score (GCS) in surviving patients than in non-survivors (Table 1).

**Table 1.** Comparison of clinical and laboratory variables between survivors and non-survivors with ALF.

| Variables | Survivors (391)<br>Mean ± SD/Median (IQR) | Non-survivors (249)<br>Mean ± SD/Median (IQR) | p value |
| --- | --- | --- | --- |
| Age (years) | 57 (43 – 69) | 53 (43 – 68) | 0.38 |
| Male Sex, n (%) | 204 (52.17) | 148 (59.44) | **0.072** |
| ICU Length of stay (days) | 12.19 ± 15.99 | 7.15 ± 6.30 | **<0.001** |
| SOFA | 7.05 ± 3.75 | 8.70 ± 4.04 | **<0.001** |
| King's College Criteria (KCC) | 1.00 (0.00 – 1.00) | 1.00 (0.00 – 1.50) | **<0.001** |
| Alanine Aminotransferase (U/L) | 2082 ± 3187 | 1344 ± 2031 | **0.003** |
| Aspartate Transaminase (U/L) | 2741 ± 4081 | 2293 ± 3237 | 0.180 |
| Alkaline Phosphatase (U/L) | 104 (71 – 161) | 124 (82 – 178) | **0.007** |
| Bilirubin (mg/dL) | 2.00 (0.90 – 5.45) | 2.80 (1.00 – 8.90) | **0.016** |
| Albumin (g/dL) | 3.04 ± 0.65 | 2.80 ± 0.54 | **<0.001** |
| INR | 2.66 ± 2.47 | 2.83 ± 2.47 | 0.411 |
| Urea (mg/dL) | 37.80 ± 28.94 | 43.12 ± 27.76 | **0.022** |
| Creatinine (mg/dL) | 2.21 ± 1.83 | 2.46 ± 1.91 | 0.093 |
| Sodium (mEq/L) | 139.99 ± 5.43 | 140.10 ± 5.94 | 0.811 |
| Chloride (mEq/L) | 107.28 ± 7.04 | 106.07 ± 7.29 | **0.038** |
| Phosphate (mg/dL) | 4.37 ± 2.44 | 5.33 ± 2.24 | **<0.001** |
| Bicarbonate (mEq/L) | 22.55 ± 4.70 | 22.36 ± 5.18 | 0.984 |
| Lactate (mmol/L) | 3.10 (1.90 - 5.55) | 5.05 (2.80 - 8.80) | **<0.001** |
| Arterial blood pH | 7.42 ± 0.07 | 7.39 ± 0.11 | **0.004** |
| Glucose (mg/dL) | 177.36 ± 80.81 | 202.65 ± 105.80 | **0.001** |
| Haemoglobin (mg/dL) | 11.65 ± 2.15 | 11.32 ± 2.13 | 0.058 |
| Platelet count (x 1000/μl) | 198 ± 118 | 180 ± 108 | 0.061 |
| White Blood Count (x 1000/μl) | 14.31 ± 9.33 | 14.86 ± 8.10 | 0.453 |
| Temperature ($^{\circ}$C) | 36.88 ± 1.22 | 36.69 ± 1.63 | 0.089 |
| Heart Rate (beat/min) | 94 ± 18 | 95 ± 18 | 0.522 |
| Mean Blood Pressure (mmHg) | 81.07 ± 12.00 | 78.02 ± 11.95 | **0.002** |
| Respiratory Rate (breath/min) | 20.55 ± 4.97 | 21.24 ± 4.92 | 0.084 |
| SpO<sub>2</sub> (%) | 96.74 ± 2.97 | 95.84 ± 4.71 | **0.003** |
| Glasgow Coma Score (GCS) | 9.27 ± 5.11 | 7.62 ± 4.81 | **<0.001** |

The data are expressed as the mean ± standard deviation or median (interquartile range) depending on the type of variable. SOFA; Sequential Organ Failure Assessment score, INR; International Normalized Ratio, SpO<sub>2</sub>; Oxygen Saturation, KCC; King's College Criteria. Either t tests or Mann–Whitney U tests were used for statistical analysis based on the normality of the data.

### Differences in Parenclitic network indices between ICU survivors and non-survivors

There was an overall greater correlation between variables in the survivors than in the non-survivors (Figure 1). Detection of clusters within the networks using the k-clique percolation method showed observable differences in the pattern of network clusters between the groups with the optimum k number of 5. Specifically, a cluster of liver function-related biomarkers was found only in survivors. Additionally, arterial blood pH was strongly correlated with kidney function markers (serum creatinine) in survivors (Figure 1), while in non-survivors, arterial pH was correlated with oxygen saturation and the respiratory rate (Figure 2). In addition to the correlation map, we mapped the parenclitic network of individual patients based on the deviation of pairs of physiological variables from the reference model. In general, the significant differences in the parenclitic deviation along all physiological axes were significantly greater in non-survivors than in survivors, except along the chloride-bicarbonate and creatinine-alkaline phosphatase axes (Table 2).

**Figure 1.**
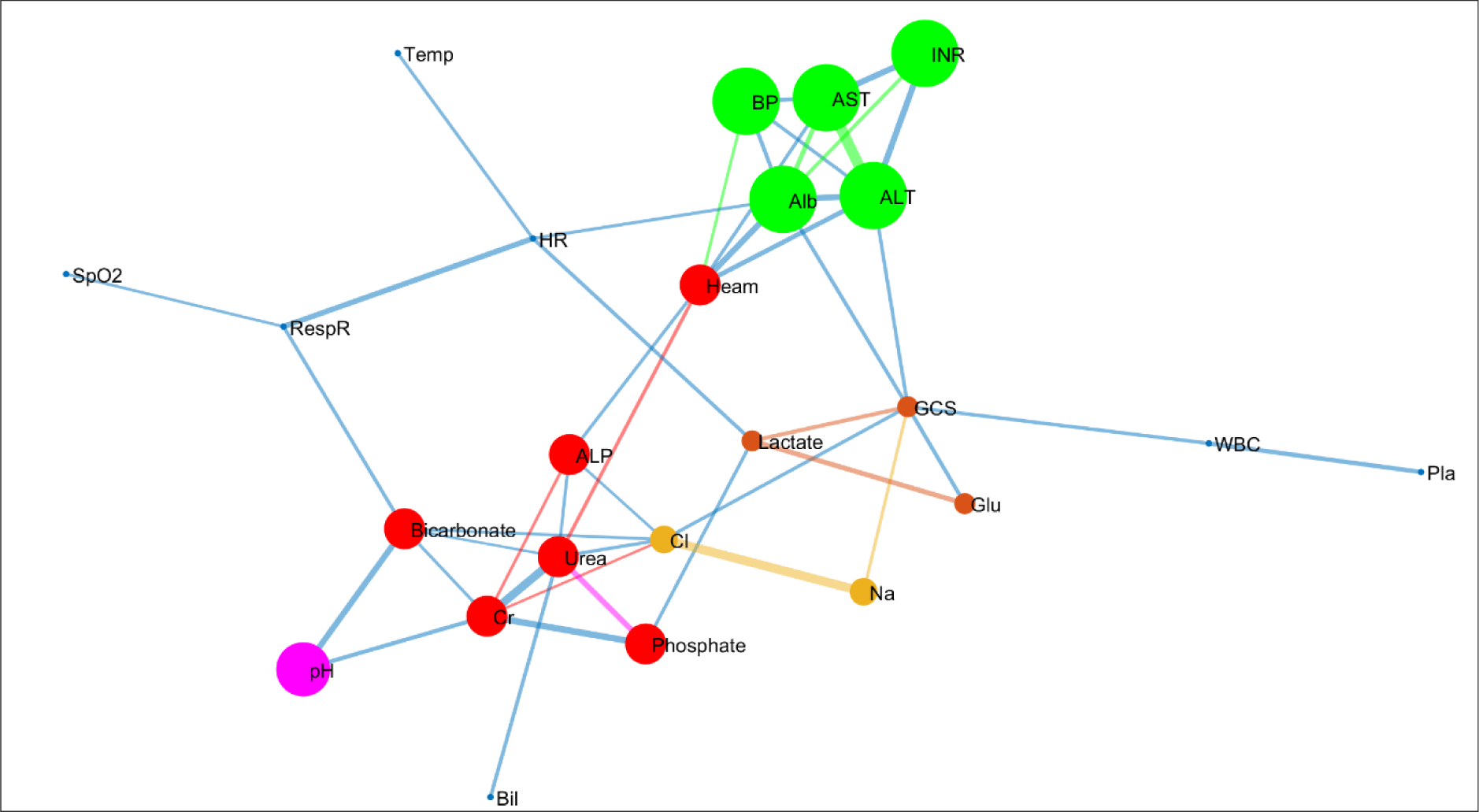
Network map of clinical and laboratory variables showing correlations and k-clique percolation clusters of patients with acute liver failure who survived their ICU stay (optimized k - clique size = 3). Cl, chloride; AST, aspartate transaminase; ALT, alanine aminotransferase; GCS, Glasgow Coma Score; Bil, total bilirubin; ALP, alkaline phosphatase; Cr, serum creatinine; Na, serum sodium; Glu, blood glucose; HR, heart rate; RespR, respiratory rate; Temp, temperature.

**Figure 2.**
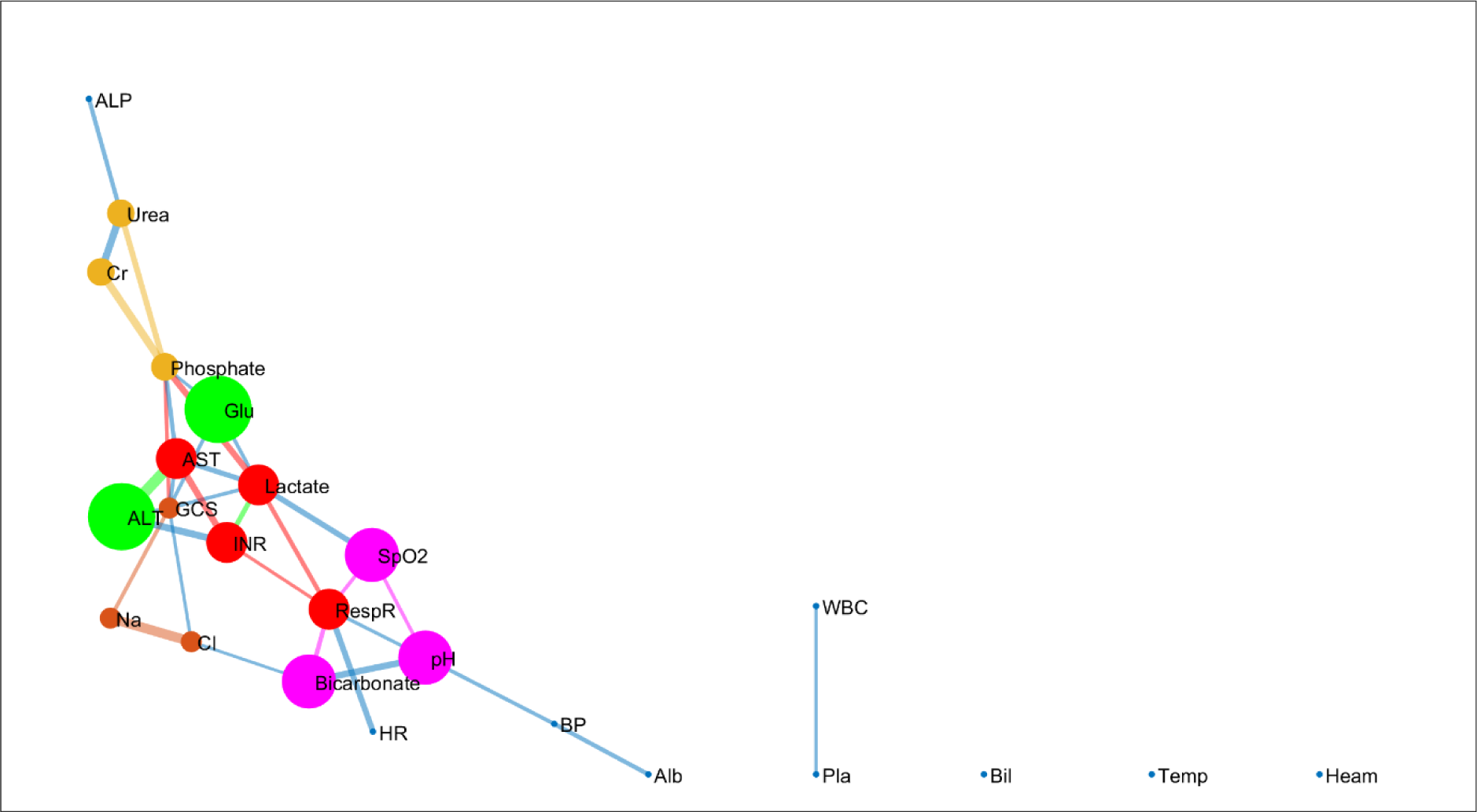
Network map of clinical and laboratory variables showing correlations and K-Clique percolation communities of patients with acute liver failure who did not survive their ICU stay (Optimized k - clique size = 3). Cl, chloride; AST, aspartate transaminase; ALT, alanine aminotransferase; GCS, Glasgow Coma Score; Bil, total bilirubin; ALP, alkaline phosphatase; Cr, serum creatinine; Na, serum sodium; Glu, blood glucose; HR, heart rate; RespR, respiratory rate; Temp, temperature.

**Table 2.**
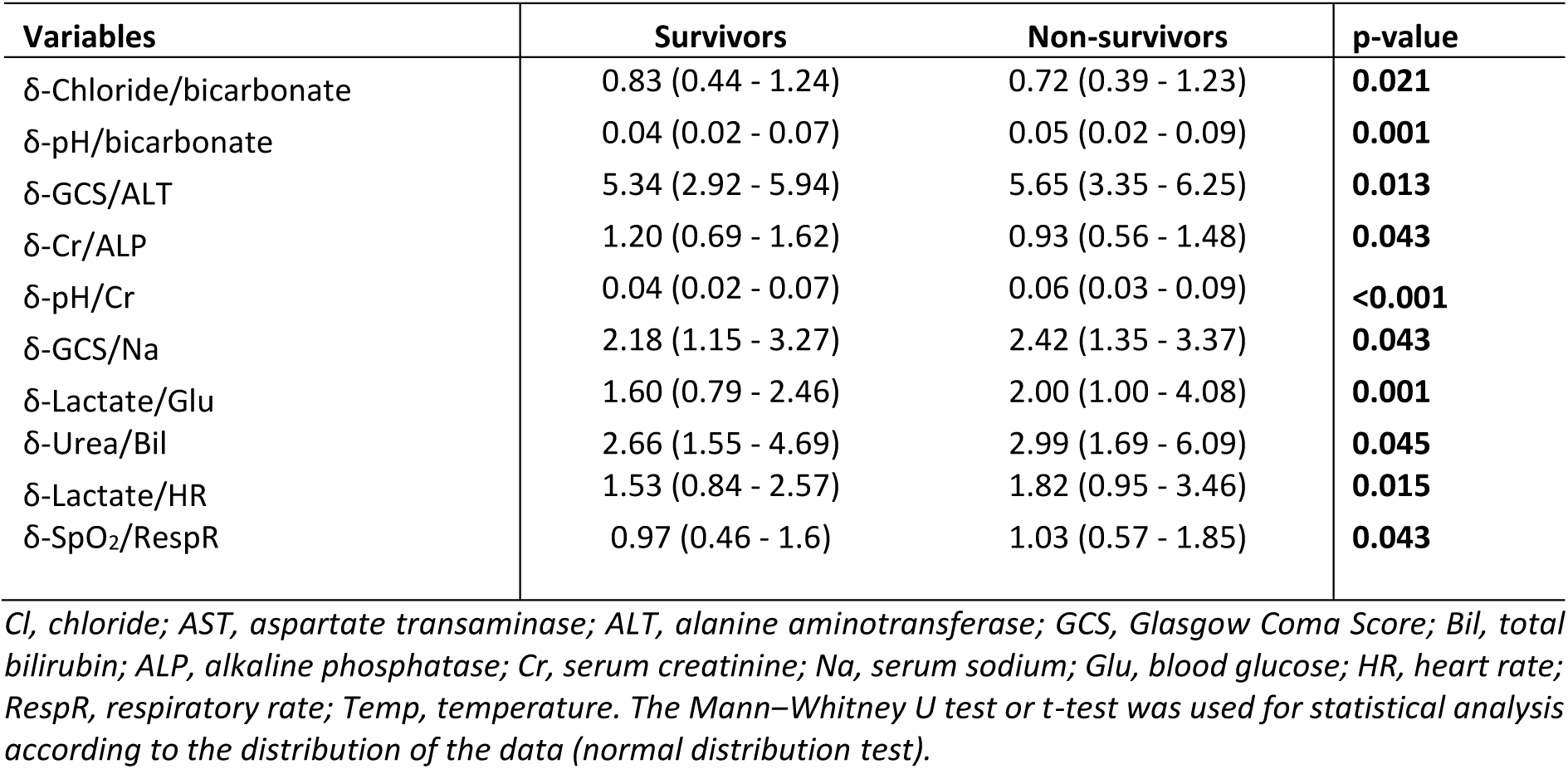
Comparison of pulmonary deviations (δ) between survivors and non-survivors in patients with ALF.

### Parenclitic deviations predict survival

Parenclitic deviations along the pH-bicarbonate, pH-creatinine, lactate-glucose, lactate-heart rate, and SpO_2_-respiratory rate axes were significantly linked with an increased risk of 28-day ICU mortality according to univariate Cox regression analysis. Specifically, each unit of deviation along the pH-bicarbonate and pH-creatinine axes was associated with greater than 37% and 36% increases, respectively, in the risk of ICU mortality. Additionally, unit deviations along the lactate-glucose, lactate-heart rate, and SpO_2_-respiratory rate axes were linked to approximately 40%, 36%, and 21% increases in the risk of 28-day mortality in the ICU, respectively (Table 3).

**Table 3.** Univariate Cox regression analysis of parenclitic indices based on ICU survival and 28-day follow-up.

| Variables | $\beta$ | SEM | Hazard Ratio (95% CI) | p value |
| --- | --- | --- | --- | --- |
| δ-Chloride/bicarbonate | 0.079 | 0.094 | 1.08 (0.90 – 1.30) | 0.402 |
| δ-pH/bicarbonate | 0.317 | 0.082 | 1.37 (1.17 – 1.61) | **<0.001** |
| δ-GCS/ALT | 0.078 | 0.127 | 1.08 (0.84 – 1.39) | 0.539 |
| δ-Cr/ALP | 0.119 | 0.085 | 1.13 (0.95 – 1.33) | 0.162 |
| δ-pH/Cr | 0.308 | 0.078 | 1.36 (1.17 – 1.59) | **<0.001** |
| δ-GCS/Na | - 0.066 | 0.107 | 0.94 (0.76 – 1.15) | 0.536 |
| δ-Lactate/Glu | 0.334 | 0.076 | 1.40 (1.20 – 1.62) | **<0.001** |
| δ-Urea/Bil | 0.088 | 0.073 | 1.09 (0.95 – 1.26) | 0.228 |
| δ-Lactate/HR | 0.307 | 0.077 | 1.36 (1.17 – 1.58) | **<0.001** |
| δ-SpO<sub>2</sub>/RespR | 0.190 | 0.070 | 1.21 (1.05 – 1.39) | **0.007** |

### Parenclitic deviations predict survival independent of SOFA score and the King’s College *Criteria*

Multivariate Cox regression analyses were performed to assess whether the significantly predictive network indices were independent of the severity of ALF, as measured by the SOFA score and KCC. SOFA score and KCC were initially assessed for their predictive value and were both found to be individual predictors of 28-day mortality in this study population (hazard ratio, 95% CI = 1.04, 1.01 – 1.08, p = 0.014 and 1.18, 1.02 – 1.34, p = 0.034, respectively). Accordingly, multivariate analysis revealed that physiological deviations along the blood pH-bicarbonate (hazard ratio, 95% CI = 1.32, 1.11 – 1.56), arterial pH-serum creatinine (hazard ratio, 95% CI = 1.30, 1.11 – 1.52), lactate-glucose (hazard ratio, 95% CI = 1.34, 1.15 – 1.57), lactate-heart rate (hazard ratio, 95% CI = 1.32, 1.14 – 1.54) and SpO_2_-respiratory rate (hazard ratio, 95% CI = 1.19, 1.03 – 1.36) axes predict 28-day mortality independent of the SOFA score and King’s College Criteria (Table 4).

**Table 4.** Multivariate Cox regression of parenclitic indices vs SOFA and King’s College Criteria (KCC) in predicting survival in ALF patients admitted to the ICU.

| Variables | $\beta$ | SEM | Hazard Ratio (95% CI) | p value |
| --- | --- | --- | --- | --- |
| δ-pH/bicarbonate | 0.275 | 0.086 | 1.32(1.11 – 1.56) | **0.001** |
| SOFA | 0.053 | 0.022 | 1.06(1.01 – 1.10) | **0.017** |
| KCC | 0.092 | 0.107 | 1.10(0.89 – 1.35) | 0.391 |
| δ-pH/Cr | 0.259 | 0.080 | 1.30(1.11 – 1.52) | **0.001** |
| SOFA | 0.050 | 0.023 | 1.05(1.01 – 1.10) | **0.027** |
| KCC | 0.098 | 0.107 | 1.02(0.82 – 1.26) | 0.362 |
| δ-Lactate/Glu | 0.295 | 0.079 | 1.34(1.15 – 1.57) | **<0.001** |
| SOFA | 0.032 | 0.023 | 1.03(0.99 – 1.08) | 0.164 |
| KCC | 0.113 | 0.107 | 1.10(0.89 – 1.36) | 0.289 |
| δ-Lactate/HR | 0.281 | 0.078 | 1.32(1.14 – 1.54) | **<0.001** |
| SOFA | 0.033 | 0.023 | 1.03(0.99 – 1.08) | 0.159 |
| KCC | 0.133 | 0.106 | 1.14(0.93 – 1.41) | 0.211 |
| δ-SpO<sub>2</sub>/RespR | 0.171 | 0.071 | 1.19(1.03 – 1.36) | **0.016** |
| SOFA | 0.027 | 0.021 | 1.03(0.99 – 1.07) | 0.188 |
| KCC | 0.082 | 0.098 | 1.09(0.9 – 1.31) | 0.402 |

SOFA; Sequential Organ Failure Assessment score, Cr; Serum creatinine, Glu; Blood glucose, HR; Heart rate, SEM; Standard error of the mean, CI; Confidence interval

### Principal component analysis

A total of 9 principal components had eigenvalues >1 and were included in further analysis (**Error! Reference source not found.**). Of these, only principal components 1, 3, and 7 were significantly linked with mortality according to univariate Cox regression analysis (**Error! Reference source not found.**). Furthermore, multivariate Cox regression analysis revealed that each unit increase in PC-1 and decrease in PC-3 were associated with 28% and 26% increases, respectively, in the risk of mortality in ALF patients (Table 5). PC-1 identified *ALT, AST,* and *INR,* while PC-2 identified *albumin*, *mean blood pressure,* and *haemoglobin* as clusters for dimension reduction, as shown in **Error! Reference source not found.**.

**Table 5.** Multivariate Cox regression of principal components vs SOFA score in predicting survival in ALF patients admitted to the ICU.

| Variables | $\beta$ | SEM | Hazard Ratio (95% CI) | p-value |
| --- | --- | --- | --- | --- |
| PCA-1 | 0.248 | 0.091 | 1.28 (1.07 - 1.53) | **0.006** |
| SOFA | 0.059 | 0.033 | 1.06 (1.00 - 1.13) | 0.068 |
| KCC | 0.180 | 0.145 | 1.20 (0.90 - 1.59) | 0.213 |
| PCA-3 | -0.300 | 0.098 | 0.74 (0.61 - 0.90) | **0.002** |
| SOFA | 0.042 | 0.034 | 1.04 (0.98 - 1.11) | 0.209 |
| KCC | 0.297 | 0.149 | 1.35 (1.01 - 1.80) | **0.047** |
| PCA-7 | 0.164 | 0.105 | 1.18 (0.96 - 1.45) | 0.117 |
| SOFA | 0.054 | 0.033 | 1.06 (0.99 - 1.13) | 0.103 |
| KCC | 0.171 | 0.152 | 1.19 (0.88 - 1.60) | 0.261 |

SEM; standard error of the mean, CI; confidence interval, SOFA; Sequential Organ Failure Assessment score.

### Parenclitic network indices improve the predictive value of SOFA score

Based on the area under the ROC curve, the addition of both parenclitic indices and principal components to the SOFA score significantly improved its predictive value, as shown by the percentage increase in the area under the curve (AUC) as well as the relatively lower Brier scores of the composite scores compared with those of the SOFA score alone (Table 6). The results from the IDI and NRI analyses showed that the addition of the parenclitic indices and principal components significantly improved the prognostic performance of the SOFA score. With regard to the IDI and NRI, the addition of principal component 1 showed the greatest reduction (IDI = 15.1% and NRI = 73.2%) in overall predictive error compared with the use of SOFA alone (Table 7).

**Table 6.** Area under the ROC curves, sensitivity, specificity, positive predictive value (PPV), negative predictive value (NPV), and Brier score of parenclitic indices and principal components in combination with the SOFA score compared with the SOFA score alone.

| Variables | AUC | p-value | Cut-Off | Sensitivity | Specificity | % AUC increase | PPV | NPV | Brier Score |
| --- | --- | --- | --- | --- | --- | --- | --- | --- | --- |
| SOFA | 0.617 | **<0.001** | 6.5 | 0.683 | 0.56 | - | 0.497 | 0.735 | 0.1294 |
| δ-pH/Carbonate + SOFA | 0.658 | **<0.001** | 0.725 | 0.606 | 0.614 | 6.65 | 0.5 | 0.71 | 0.1268 |
| δ-pH/Creatinine + SOFA | 0.633 | **<0.001** | 0.695 | 0.601 | 0.592 | 2.59 | 0.484 | 0.7 | 0.1276 |
| δ-Lactate/HR + SOFA | 0.646 | **<0.001** | 0.4954 | 0.657 | 0.626 | 4.7 | 0.528 | 0.741 | 0.1231 |
| δ-Lactate/Glu + SOFA | 0.652 | **<0.001** | 0.4902 | 0.684 | 0.641 | 5.67 | 0.548 | 0.761 | 0.1232 |
| δ-SpO<sub>2</sub>/RespR + SOFA | 0.654 | **<0.001** | 0.6461 | 0.623 | 0.619 | 6 | 0.51 | 0.721 | 0.1226 |
| PC-1 + SOFA | 0.754 | **<0.001** | 0.6422 | 0.715 | 0.714 | 22.2 | 0.614 | 0.797 | 0.1428 |
| PC-3 + SOFA | 0.712 | **<0.001** | 0.5692 | 0.692 | 0.667 | 15.4 | 0.57 | 0.773 | 0.1344 |

**Table 7.** The SOFA score improved the prognosis due to the addition of parenclitic indices and principal components.

| Variables | IDI | p-value | NRI | p-value |
| --- | --- | --- | --- | --- |
| δ-pH/Bicarbonate + SOFA | 0.0539 | **<0.001** | 0.3928 | **<0.001** |
| δ-pH/Creatinine + SOFA | 0.0521 | **<0.001** | 0.3939 | **<0.001** |
| δ-Lactate/Heart Rate + SOFA | 0.0464 | **<0.001** | 0.3790 | **<0.001** |
| δ-Lactate/Glucose + SOFA | 0.0401 | **<0.001** | 0.3641 | **<0.001** |
| δ-SpO<sub>2</sub>/Respiratory Rate + SOFA | 0.0406 | **<0.001** | 0.3723 | **<0.001** |
| PC-1 + SOFA | 0.0061 | 0.2082 | 0.1755 | 0.1760 |

IDI: integrated discrimination improvement, NRI: net reclassification indices.

Additionally, Kaplan–Meier survival curves (with the Mantel–Cox test) showed that compared with the SOFA score alone, the ROC cut-off of the composite score significantly differentiated between ICU-admitted ALF patients who survived and those who did not. Specifically, the cut-off values of the composite scores, including the SOFA score and independent predictive platelet deviations (δ-pH/bicarbonate, p < 0.001; δ-pH/creatinine, p = 0.005; δ-lactate/glucose, p = 0.041; δ-lactate/heart rate, p = 0.001), significantly classified survivors and non-survivors (Figure 3 and 4).

**Figure 3.**
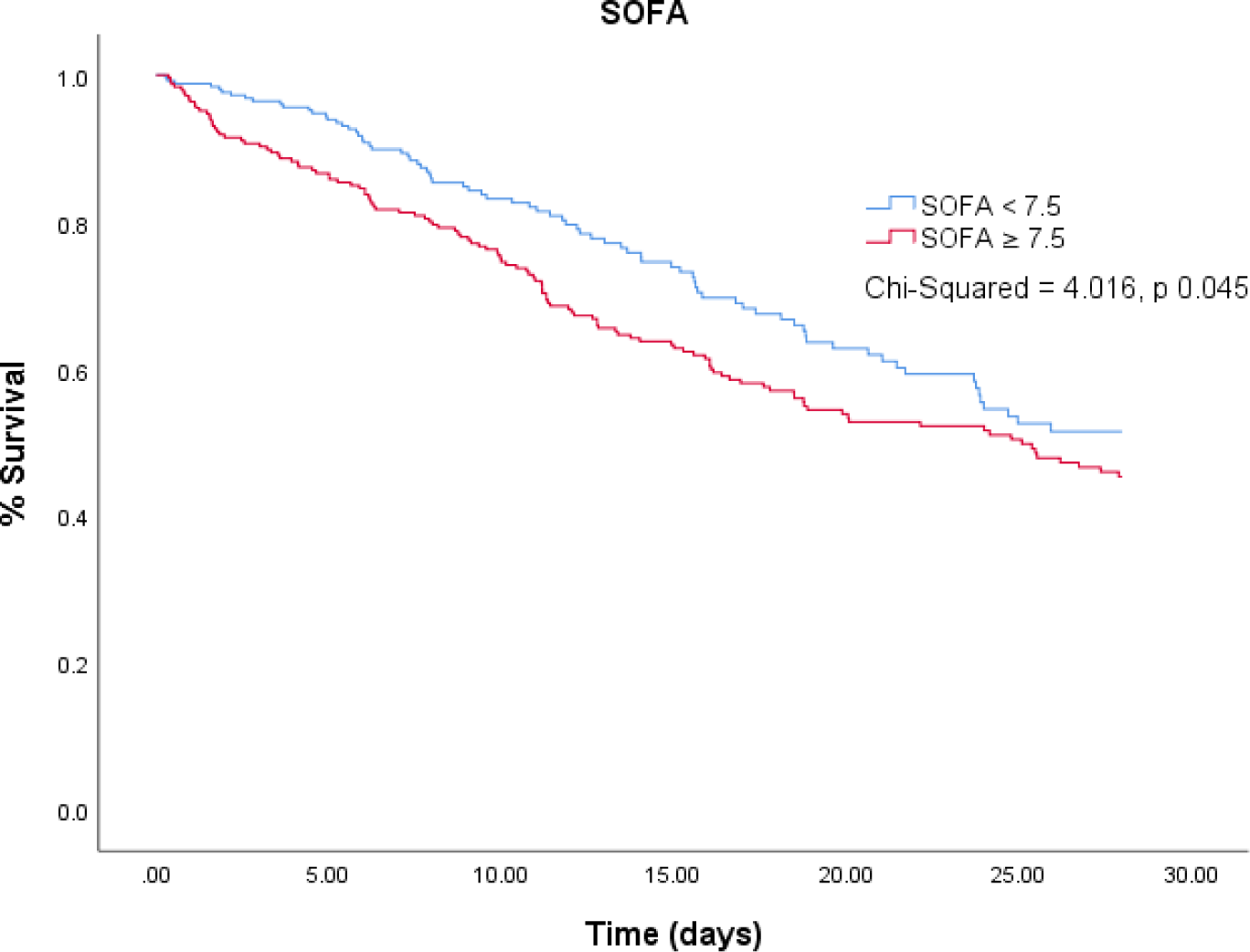
Kaplan–Meier graphs of patients with acute liver failure admitted to the intensive care unit who survived and those who did not survive after 28 days according to SOFA cut-off scores.

**Figure 4.**
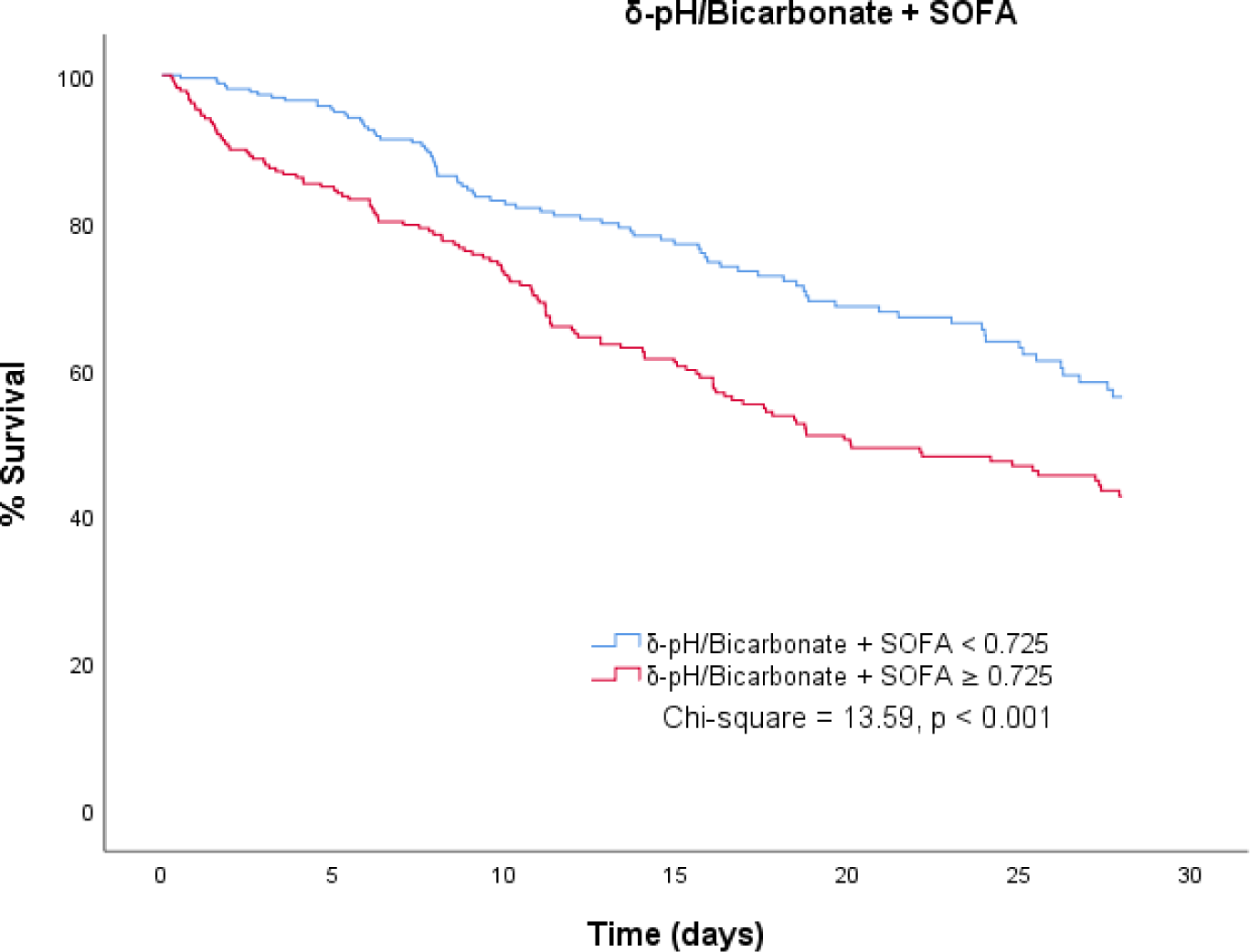
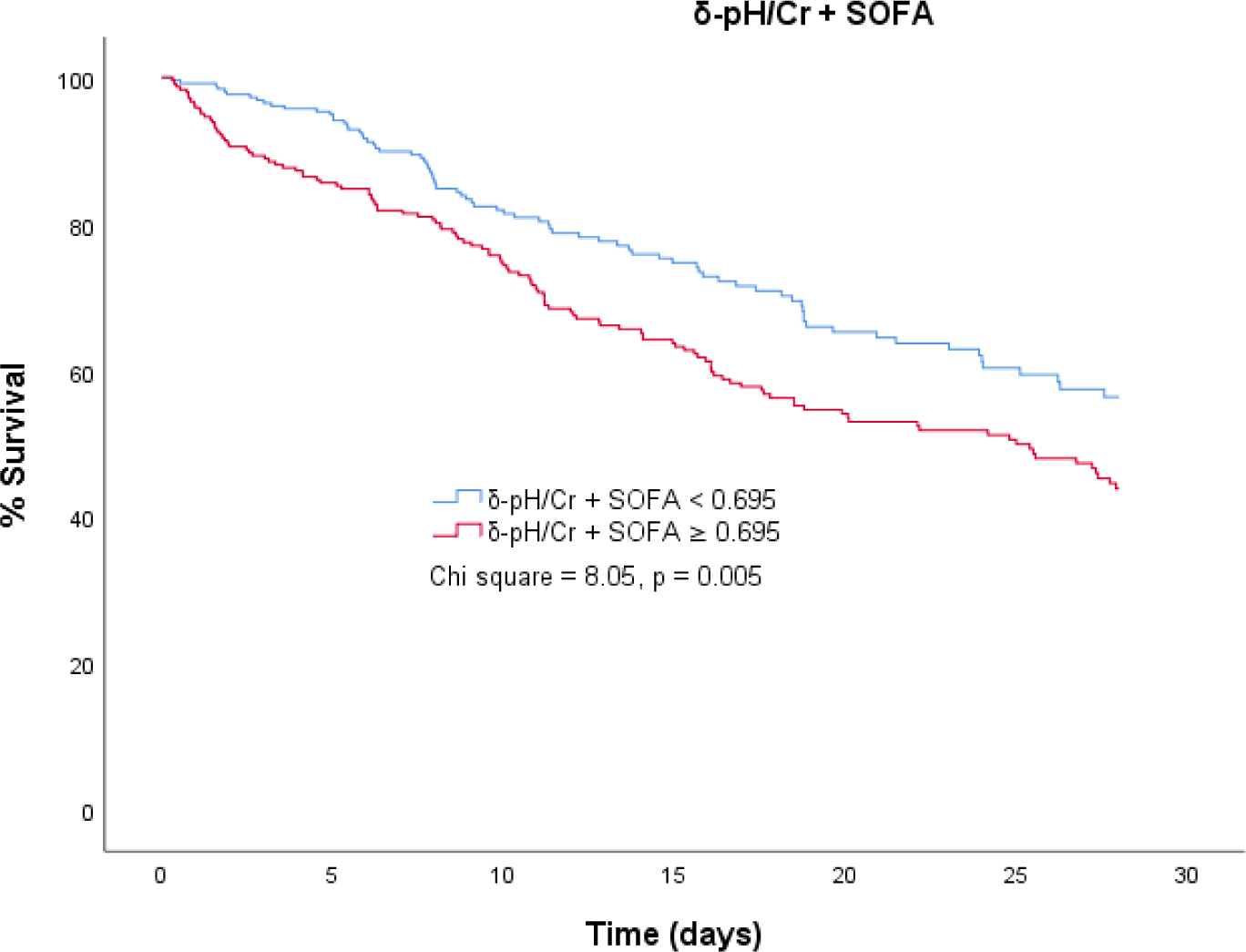
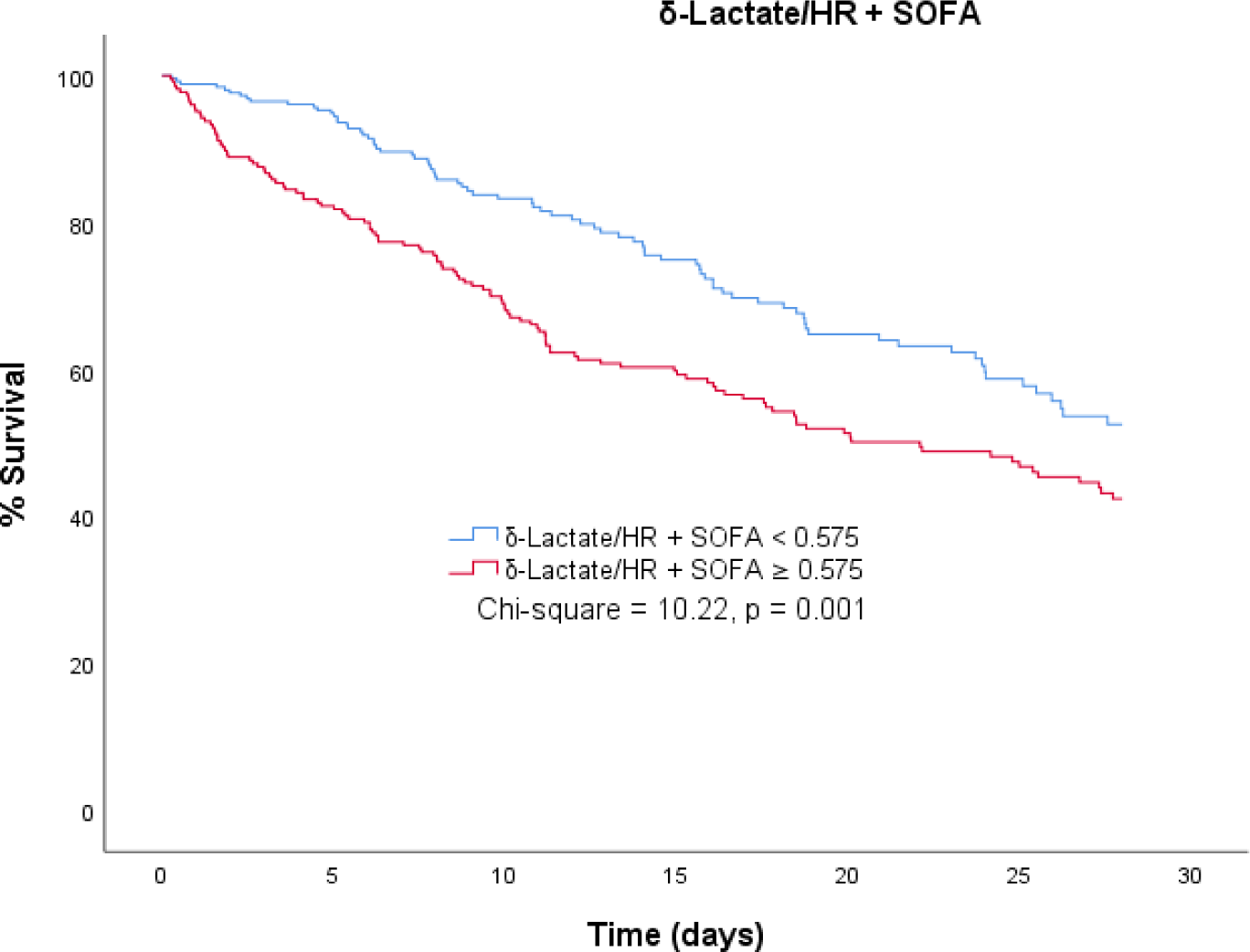
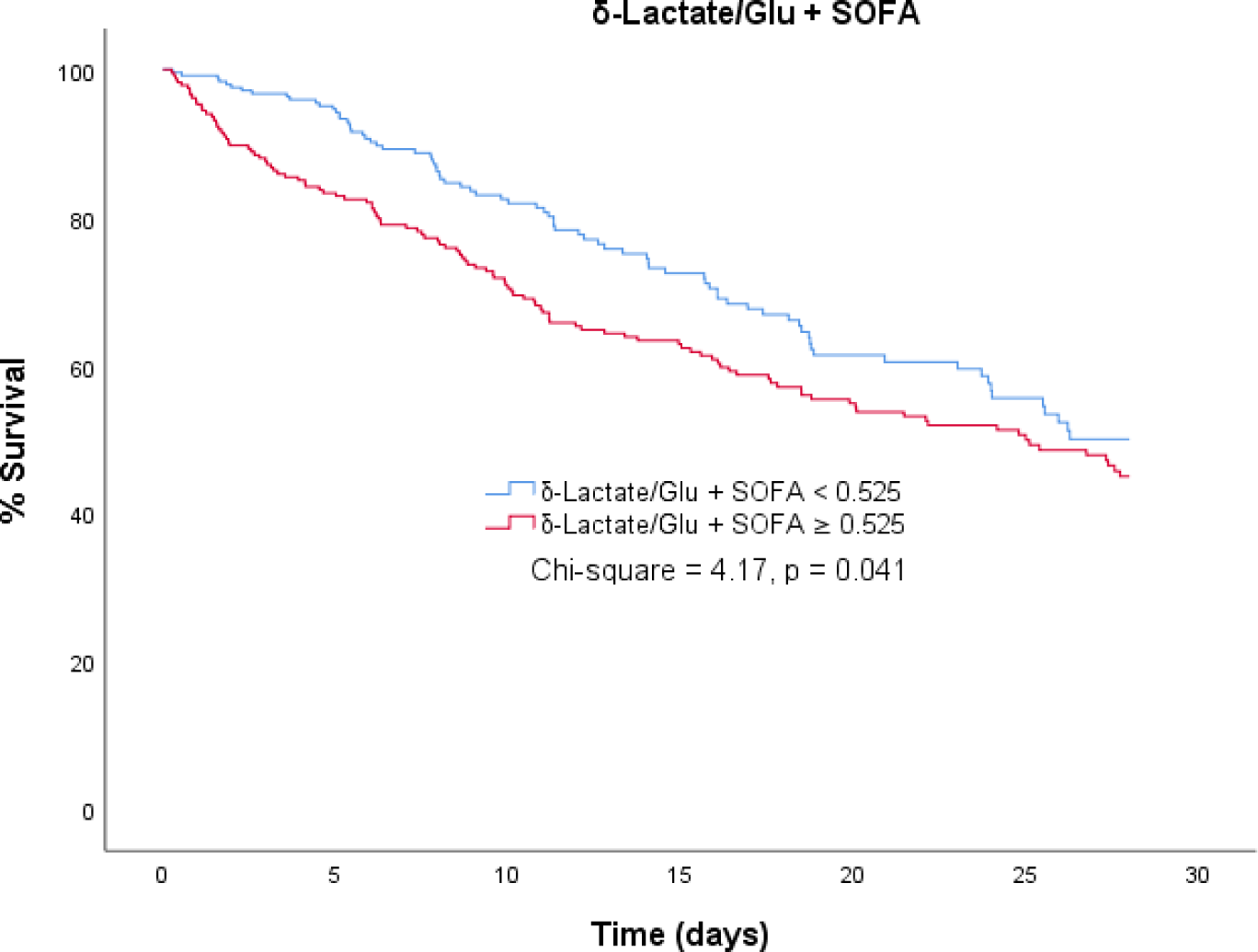
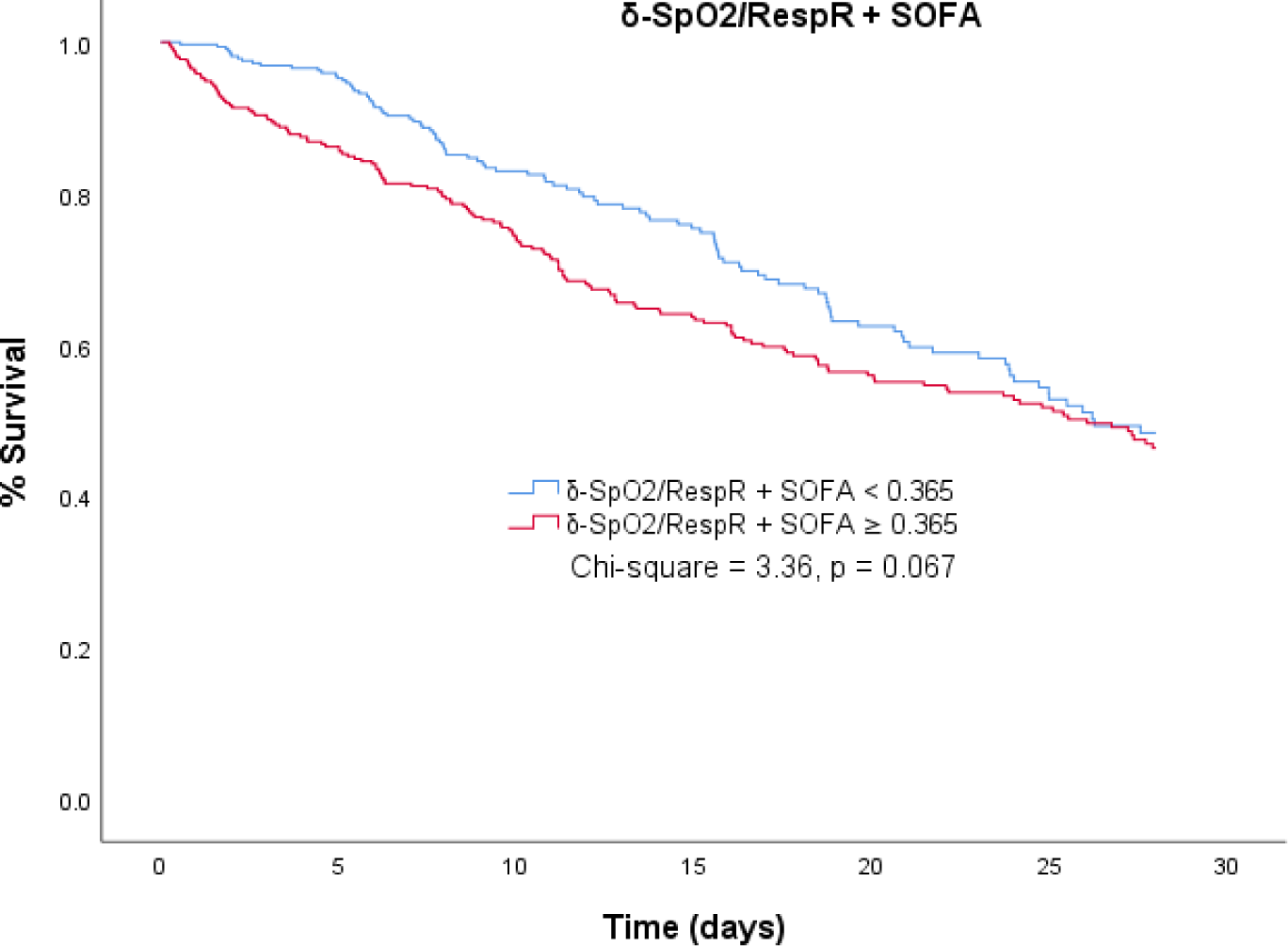
Kaplan–Meier graphs of patients with acute liver failure admitted to the intensive care unit who survived and those who did not survive after 28 days, as classified by the cut-offs of the composite scores from the combination of SOFA with the parenclitic deviations along the pH-bicarbonate (pH-CO3), pH-creatinine (pH-Cr), lactate-heart rate (lactate-HR), lactate-glucose (lactate-Glu), and oxygen saturation-respiratory rate axes.

## Discussion

The liver serves as a major hub in the physiological network, and acute dysfunction is associated with mortality in many patients. Those who survive ALF may have adapted compensatory mechanisms that enhance survival. However, identifying these mechanisms necessitates a holistic or network approach. In this study, we employed parenclitic network mapping to demonstrate the prognostic value of this network approach in predicting survival among ICU-admitted patients with paracetamol-induced ALF. The results revealed that parenclitic deviations across different physiological axes can predict survival in this patient population independent of the current clinical prognostic factors (SOFA score and KCC). Additionally, compared with the SOFA score alone, the combination of the independent survival predictors with the SOFA score resulted in a greater than 30% reduction in classification error. This is the first study, to the best of our knowledge, to apply integrative network mapping in the prediction of survival in ICU-admitted ALF patients.

In terms of the correlation network, ALF patients who did not survive in the ICU for at least 28 days were found to have overall lower organ system connectivity than survivors. Furthermore, network community detection also revealed a marked difference in network structure between the survivors and non-survivors characterized by a significant difference in organ system clustering (community formation). Generally, a community in a network defines a subpopulation of nodes (organ systems) that are more closely linked (clustered) than other nodes outside of the community [38–40]. As expected, the variables associated with liver function (e.g., ALT, AST, INR, and Alb) were relatively more clustered in survivors than in non-survivors (green nodes in Figure 1 *vs* Figure 2). This finding is in line with the results of the principal component analysis, as PC-1 in this analysis also revealed ALT, AST and INR in the same cluster. According to survival analysis, this cluster (PC-1), could predict mortality independently of the SOFA score and KCC.

In addition to the liver function community in the correlation network, arterial pH was also found to cluster with serum creatinine and bicarbonate in survivors compared with non-survivors, where it clustered with respiratory nodes (SpO_2_, respiratory rate) and bicarbonate (purple nodes in Figure 1 and Figure 2). Thus, physiologically different compensatory mechanisms for acid‒base balance between the groups were inferred. Essentially, it appears that the regulation of arterial pH was more closely linked with renal function in survivors, while in non-survivors, this function was mainly modulated by respiratory compensation. The role of the liver in acid‒base homeostasis via the regulation of systemic clearance of lactic acid and urea is generally dysregulated in liver disease [41]. This means that in liver disease, the role is generally shifted to classical regulatory pathways involving the respiratory and renal systems. In the context of ALF, acid-base disequilibrium has been previously shown to be driven mainly by a systemic increase in lactic acid, especially due to overproduction in peripheral organs [42]. Generally, the kidney plays a protective role in lactic acidosis through a pH-dependent increase in the rate of clearance of systemic lactic acid [43,44]. Indeed, lactic acidosis is significantly linked with poorer prognosis in patients with ALF [45–50]. Although ALF patients typically exhibit markedly increased lactate levels, frequently, there is no evident acid-base imbalance due to compensatory hypoalbuminemia alkalosis [51]. In our study, arterial pH was only slightly lower in non-survivors with ALF than in survivors (Table 1). It appears that sufficient renal function may be associated with improved survival in ALF patients admitted to the ICU, especially since the non-survivors in this study showed significantly greater deviation in the pH-creatinine axis (δ-pH/creatinine, Tables 2-4). In line with this observation, δ-pH/creatinine was an independent prognostic factor for mortality, indicating the importance of this axis in the survival of critically ill patients with ALF.

Aside from connectivity and organ system community structure, overall parenclitic deviations of non-survivors were found to be significantly greater than those of survivors. For instance, non-survivors have greater deviation along the pH-bicarbonate, pH-creatinine, lactate-glucose, lactate-heart rate, and SpO_2_-respiratory rate axes, which are independently associated with survival. This translates to significantly reduced physiological coupling between these pairs of variables (i.e., higher parenclitic deviation means higher deviation from the regression line between two biomarkers in the reference population). Specifically, most of the physiological disconnections observed in non-survivors are associated with acid-base homeostasis (i.e., arterial pH, serum bicarbonate level, and serum lactate). These findings support the importance of acid‒base compensatory mechanisms in the prognosis of patients with ALF and critically ill patients [52]. Importantly, these network deviations predicted mortality even though there was only a slight difference in arterial pH between the patient groups (7.42 ± 0.07 versus 7.39 ± 0.11). Thus, considering the physiological network context rather than the individual isolated organ system is important for clinical management and prognosis. Additionally, deviation along the SpO_2_-respiratory rate axes predicted survival independent of SOFA score and KCC. SpO_2_ estimates the percentage of oxyhaemoglobin relative to the total blood haemoglobin and reflects cardiopulmonary efficiency in terms of systemic oxygen transportation. Although multiple cardiorespiratory factors contribute to oxygen saturation dynamics [8], SpO_2_ and the respiratory rate have been shown to be physiologically inversely correlated [53]. Interestingly, there was a significantly greater mean baseline SpO_2_ and lower respiratory rate in ALF patients who survived than in non-survivors (). Thus, the loss of correlation and coordination between these two variables may be interpreted as a patient’s loss of adaptability and reduced ability to maintain systemic oxygen levels. Indeed, a previous study by Mower et al., showed a poor correlation between oxygen saturation and respiratory rate in approximately 15,000 patients admitted to the ICU for various reasons. However, the authors did not assess the relationship with patient survival [54].

Indeed, the results of this study corroborate previous research in the field, where changes in organ system connectivity measured using network analysis and other methods were shown to improve traditional scoring systems for predicting patient outcomes in patients with cirrhosis or even detecting subgroups of patients who may respond to therapy (e.g., targeted albumin therapy)[17,18,55,56]. Specifically, a previous study using parenclitic network analysis showed reduced organ system connectivity (based on population-level correlation network mapping) in patients with cirrhosis who did not survive compared with survivors [18]. This corroborated the findings of a previous study in patients with cirrhosis referred for formal clinical assessment of hepatic encephalopathy, with similar findings showing that organ system connectivity was relatively lower in patients who did not survive after 12 months of follow-up [17]. However, the reduction observed in the patients with decompensated cirrhosis was greater in magnitude than that in the patients with ALF among the non-survivors. This may be due to the significantly different time course of development or clinical history of these diseases, whereby cirrhosis often develops over decades, while ALF progresses more rapidly over the course of a few weeks or days in otherwise healthy individuals [57,58]. Thus, while chronic cirrhosis probably affects the overall physiology of patients, culminating in a general loss of organ system coupling, ALF is likely linked to a shift in compensatory mechanisms to counter abrupt and rapidly deteriorating liver function. Regardless of the time course of liver failure, a physiological network approach appears to provide information not currently offered by existing clinical criteria (e.g., SOFA). This could open new avenues for improved prognostication and the discovery of novel personalized therapies based on individual networks in future investigations.

To further verify the results of the network analysis, especially network community detection, we performed principal component analysis to assess whether similar variables are combined within each component and whether these components can also predict survival in ALF patients. While principal component analysis is useful for dimension reduction in multivariate datasets, it is limited in scope compared with network mapping (e.g., correlation or parenclitic). Thus, while it could be used for validation, it does not suffice as a viable replacement for network analysis. This is due to several inherent limitations of principal component analysis. First, PCA is prone to the loss of crucial information and oversimplicity resulting from the reduction of dataset dimensions [59]. Importantly, because PCA is based on linear correlations between variables in the overall population, compared with Parenclitic network analysis, PCA does not consider the possibility that correlations may be different between subgroups (e.g., survivors and non-survivors, healthy individuals and diseased individuals) within the study population.

Indeed, the principal components related to liver function (component 1; AST, ALT, and INR), hemodynamic function (component 4; serum albumin, mean blood pressure, and haemoglobin), and metabolic function (component 7; lactate, glucose, e.g., liver-dependent Cori cycle for lactate-to-glucose recycling) were found to be significantly associated with patient survival. However, only components 1 (liver function) and 3 (haemodynamic) were independent of the SOFA score and the KCC (**Error! Reference source not found.**). Essentially, component 1 captures the ALF-related rapid deterioration of liver function due to paracetamol-induced, cytochrome P450-driven toxicity, as well as increased intrahepatic glutathione depletion, oxidative stress, and mitochondrial dysfunction resulting in hepatocyte necrosis [60,61]. This finding is in line with a recent study by Yang et al., which revealed that the addition of INR (and alkaline phosphatase) could significantly improve Hy’s model for predicting patients with drug-induced liver injury (DILI) likely to develop ALF [62]. Hy’ model is a risk scoring model developed in 1968 by Hyman Zimmerman for the prediction of the progression to ALF in patients with DILI and incorporates patients’ total bilirubin, AST, and ALT [63]. Indeed, due to the relatively low specificity, other variations to the original Hy’s law and other prognostic models have been proposed and validated, including the “new Hy’s law”, which has higher specificity [64–66]. Additionally, the ratio of AST to ALT remains a popular model for the diagnosis and prognosis of various aetiologies of liver disease, including those of primary biliary, alcoholic, or viral origin [67–69].

Furthermore, our results show that hemodynamic dysregulation, represented by component 3 (serum albumin, mean blood pressure, and haemoglobin), is also significantly linked to poorer prognosis in patients with ALF. This finding is in line with previous works in the field showing the implication of haemodynamic dysregulation and instability in the prognosis of acute liver failure [70]. ALF is clinically linked to dysfunction in multiple organ systems, including hyperdynamic circulation characterized by hypotension (reduced mean arterial pressure), hyperdynamic circulation, and reduced vascular resistance [71]. Thus, a reduction in renal perfusion pressure resulting from hemodynamic dysregulation remains the key driver of renal failure, a complication observed in up to 82% of patients with ALF and linked with a significantly poor prognosis [72–75]. Indeed, the haemodynamic changes in ALF patients have been shown to correlate with the severity of liver disease and are a strong determinant of liver transplantation outcome [76].

Finally, principal component 7, comprising glucose and lactate levels, also predicted mortality, albeit not independently of patient severity score. The Cori cycle describes the conversion of muscle-generated lactate, a byproduct of anaerobic glycolysis, to glucose in the liver. Physiologically, gluconeogenesis converts lactate to glucose in the liver, usually in response to hypoglycaemia. Thus, it could be assumed that hypoglycaemia and hyperlactatemia should not coexist. However, in conditions such as paracetamol-induced ALF, which is characterized by severe hepatocyte necrosis and loss of liver function and dysfunction in the Cori cycle, these conditions may coexist [77,78]. Indeed, hyperlactatemia and hypoglycaemia have been shown to be individually associated with poorer prognosis in p-ALF patients [46].

### Limitations

One limitation of this study is the retrospective nature of this study, which is inherently linked with selection bias and a lack of some relevant variables (e.g., West Haven HE score), since the record was not specifically designed for the assessment of ALF [79]. Additionally, because the MIMIC-III dataset involves patients admitted to a single hospital serving a limited population, the results herein should be interpreted with this in mind. Another limitation of this study is related to the characteristics of the patient population, as patients were from a single centre in the USA and were from a specific region and demographic region. However, this study is based on one of the largest populations of p-ALF patients subjected to rigorous mathematical and statistical analysis.

### Conclusion

Reduced organ system connectivity and a shift from renal to respiratory compensatory mechanisms are associated with poorer prognosis in patients with ALF. This is further supported by physiological network disconnections along pH-associated axes, which predict mortality independent of SOFA score and KCC. Indeed, the strength of network analysis is the ability to assess the interactions of multiple organ systems in critically ill patients where the risk of multiple organ failure is high and similar findings continue to show significant promise [80,81]. Future studies could benefit from using a multicentre, multinational cohort of ALF patients to validate the findings herein.

## Acknowledgments

We would like to extend our gratitude to Prof Sara Montagnese for her advice regarding assessment of hepatic encephalopathy in patients with acute liver failure.

## Author Contributions

T.O., K.P.M., and A.R.M. contributed to the conceptualization. TO. contributed to clinical data extraction. T.O. and A.R.M. contributed to the formal analysis and software development. A.R.M. contributed to the supervision. T.O., A.R.M., and K.P.M. contributed to the writing, review, and editing of the manuscript. All authors contributed to the article and approved the submitted version.

## Funding statement and conflict of interest disclosure

The authors declare that the research was conducted in the absence of any commercial or financial relationships that could be construed as a potential conflict of interest.

## Data availability

Data are available upon request from the corresponding author.

## Notes

### Competing Interest Statement

The authors have declared no competing interest.

### Funding Statement

The authors received no funding to conduct this study

### Author Declarations

The data analysed in this study were sourced from the third version of the Medical Information Mart for Intensive Care (MIMIC-III) following training, application, and acquisition of the required permissions. MIMIC-III data has been deidentified in accordance with Health Insurance Portability and Accountability Act (HIPAA) standards and the project approved by the Institutional Review Boards of Beth Israel Deaconess Medical Center and the MIT (IRB protocol nos. 2001P001699/14 and 0403000206, respectively). The authors involved in data extraction completed mandatory online ethics training at MIT and were credentialled (IDs 10304625 and 48067739).

## References

1. Raith, E.P.; Udy, A.A.; Bailey, M.; McGloughlin, S.; MacIsaac, C.; Bellomo, R.; Pilcher, D.V. Prognostic Accuracy of the SOFA Score, SIRS Criteria, and qSOFA Score for In-Hospital Mortality Among Adults With Suspected Infection Admitted to the Intensive Care Unit. Jama 2017, 317, 290–300, doi:10.1001/jama.2016.20328.

2. Maslove, D.M.; Tang, B.; Shankar-Hari, M.; Lawler, P.R.; Angus, D.C.; Baillie, J.K.; Baron, R.M.; Bauer, M.; Buchman, T.G.; Calfee, C.S.;, et al. Redefining critical illness. Nature Medicine 2022, 28, 1141–1148, doi:10.1038/s41591-022-01843-x.

3. Nielsen, A.B.; Thorsen-Meyer, H.C.; Belling, K.; Nielsen, A.P.; Thomas, C.E.; Chmura, P.J.; Lademann, M.; Moseley, P.L.; Heimann, M.; Dybdahl, L.;, et al. Survival prediction in intensive-care units based on aggregation of long-term disease history and acute physiology: a retrospective study of the Danish National Patient Registry and electronic patient records. Lancet Digit Health 2019, 1, e78–e89, doi:10.1016/s2589-7500(19)30024-x.

4. Asada, T.; Aoki, Y.; Sugiyama, T.; Yamamoto, M.; Ishii, T.; Kitsuta, Y.; Nakajima, S.; Yahagi, N.; Doi, K. Organ System Network Disruption in Non-survivors of Critically Ill Patients. Critical Care Medicine 2016, 44.

5. Shashikumar, S.P.; Li, Q.; Clifford, G.D.; Nemati, S. Multiscale network representation of physiological time series for early prediction of sepsis. Physiol Meas 2017, 38, 2235–2248, doi:10.1088/1361-6579/aa9772.

6. Asada, T.; Doi, K.; Inokuchi, R.; Hayase, N.; Yamamoto, M.; Morimura, N. Organ system network analysis and biological stability in critically ill patients. Critical Care 2019, 23, 83, doi:10.1186/s13054-019-2376-y.

7. Ivanov, P.C. The New Field of Network Physiology: Building the Human Physiolome. Front Netw Physiol 2021, 1, 711778, doi:10.3389/fnetp.2021.711778.

8. Jiang, Y.; Costello, J.T.; Williams, T.B.; Panyapiean, N.; Bhogal, A.S.; Tipton, M.J.; Corbett, J.; Mani, A.R. A network physiology approach to oxygen saturation variability during normobaric hypoxia. Experimental Physiology 2021, 106, 151–159, 10.1113/EP088755.

9. Golding, P.L.; Smith, M.; Williams, R. Multisystem involvement in chronic liver disease. Studies on the incidence and pathogenesis. Am J Med 1973, 55, 772–782, doi:10.1016/0002-9343(73)90258-1.

10. Oyelade, T.; Canciani, G.; Carbone, G.; Alqahtani, J.S.; Moore, K.; Mani, A.R. Heart rate variability in patients with cirrhosis: a systematic review and meta-analysis. Physiol Meas 2021, 42, doi:10.1088/1361-6579/abf888.

11. Mani, A.R.; Montagnese, S.; Jackson, C.D.; Jenkins, C.W.; Head, I.M.; Stephens, R.C.; Moore, K.P.; Morgan, M.Y. Decreased heart rate variability in patients with cirrhosis relates to the presence and degree of hepatic encephalopathy. Am J Physiol Gastrointest Liver Physiol 2009, 296, G330–338, doi:10.1152/ajpgi.90488.2008.

12. Abid, N.-U.-H.; Mani, A.R. The mechanistic and prognostic implications of heart rate variability analysis in patients with cirrhosis. Physiological Reports 2022, 10, e15261, 10.14814/phy2.15261.

13. Mani, A.R.; Mazloom, R.; Haddadian, Z.; Montagnese, S. Body temperature fluctuation analysis in cirrhosis. Liver Int 2018, 38, 378–379, doi:10.1111/liv.13539.

14. Bottaro, M.; Abid, N.U.; El-Azizi, I.; Hallett, J.; Koranteng, A.; Formentin, C.; Montagnese, S.; Mani, A.R. Skin temperature variability is an independent predictor of survival in patients with cirrhosis. Physiol Rep 2020, 8, e14452, doi:10.14814/phy2.14452.

15. Abid, N.-U.-H.; Lum Cheng In, T.; Bottaro, M.; Shen, X.; Hernaez Sanz, I.; Yoshida, S.; Formentin, C.; Montagnese, S.; Mani, A.R. Application of short-term analysis of skin temperature variability in prediction of survival in patients with cirrhosis. Frontiers in Network Physiology 2024, 3.

16. Tan, Y.Y.; Montagnese, S.; Mani, A.R. Organ System Network Disruption Is Associated With Poor Prognosis in Patients With Chronic Liver Failure. Frontiers in Physiology 2020, 11.

17. Zhang, H.; Oyelade, T.; Moore, K.P.; Montagnese, S.; Mani, A.R. Prognosis and survival modelling in cirrhosis using parenclitic networks. Frontiers in Network Physiology 2022, 2, 833119.

18. Oyelade, T.; Forrest, E.; Moore, K.P.; O’Brien, A.; Mani, A.R. Parenclitic Network Mapping Identifies Response to Targeted Albumin Therapy in Patients Hospitalized With Decompensated Cirrhosis. Clin Transl Gastroenterol 2023, 14, e00587, doi:10.14309/ctg.0000000000000587.

19. Williams, R.; Schalm, S.W.; O’Grady, J.G. Acute liver failure: redefining the syndromes. The Lancet 1993, 342, 273–275, doi:10.1016/0140-6736(93)91818-7.

20. Bernal, W.; Auzinger, G.; Dhawan, A.; Wendon, J. Acute liver failure. The Lancet 2010, 376, 190–201, doi:10.1016/S0140-6736(10)60274-7.

21. Forns, J.; Cainzos-Achirica, M.; Hellfritzsch, M.; Morros, R.; Poblador-Plou, B.; Hallas, J.; Giner-Soriano, M.; Prados-Torres, A.; Pottegård, A.; Cortés, J.;, et al. Validity of ICD-9 and ICD-10 codes used to identify acute liver injury: A study in three European data sources. Pharmacoepidemiology and Drug Safety 2019, 28, 965–975, 10.1002/pds.4803.

22. Udo, R.; Maitland-van der Zee, A.H.; Egberts, T.C.G.; den Breeijen, J.H.; Leufkens, H.G.M.; van Solinge, W.W.; De Bruin, M.L. Validity of diagnostic codes and laboratory measurements to identify patients with idiopathic acute liver injury in a hospital database. Pharmacoepidemiology and Drug Safety 2016, 25, 21–28, 10.1002/pds.3824.

23. Teasdale, G.; Jennett, B. ASSESSMENT OF COMA AND IMPAIRED CONSCIOUSNESS: A Practical Scale. The Lancet 1974, 304, 81–84, 10.1016/S0140-6736(74)91639-0.

24. O’Grady, J.G.; Alexander, G.J.M.; Hayllar, K.M.; Williams, R. Early indicators of prognosis in fulminant hepatic failure. Gastroenterology 1989, 97, 439–445.

25. Jain, S.; Iverson, L.M. Glasgow coma scale. 2018.

26. Montagnese, S.; Russo, F.P.; Amodio, P.; Burra, P.; Gasbarrini, A.; Loguercio, C.; Marchesini, G.; Merli, M.; Ponziani, F.R.; Riggio, O.; Scarpignato, C. Hepatic encephalopathy 2018: A clinical practice guideline by the Italian Association for the Study of the Liver (AISF). Digestive and Liver Disease 2019, 51, 190–205, 10.1016/j.dld.2018.11.035.

27. Wendon, J.; Cordoba, J.; Dhawan, A.; Larsen, F.S.; Manns, M.; Nevens, F.; Samuel, D.; Simpson, K.J.; Yaron, I.; Bernardi, M. EASL Clinical Practical Guidelines on the management of acute (fulminant) liver failure. Journal of hepatology 2017, 66, 1047–1081.

28. Vincent, J.L.; Moreno, R.; Takala, J.; Willatts, S.; De Mendonça, A.; Bruining, H.; Reinhart, C.K.; Suter, P.; Thijs, L.G. The SOFA (Sepsis-related Organ Failure Assessment) score to describe organ dysfunction/failure: On behalf of the Working Group on Sepsis-Related Problems of the European Society of Intensive Care Medicine (see contributors to the project in the appendix). 1996.

29. Shirazi, A.H.; Badie Modiri, A.; Heydari, S.; Rohn, J.L.; Jafari, G.R.; Mani, A.R. Evolution of Communities in the Medical Sciences: Evidence from the Medical Words Network. PLOS ONE 2016, 11, e0167546, doi:10.1371/journal.pone.0167546.

30. Palla, G.; Derényi, I.; Farkas, I.; Vicsek, T. Uncovering the overlapping community structure of complex networks in nature and society. Nature 2005, 435, 814–818, doi:10.1038/nature03607.

31. Palla, G.; Derényi, I.; Farkas, I.; Vicsek, T. Uncovering the overlapping community structure of complex networks in nature and society. Nature 2005, 435, 814–818, doi:10.1038/nature03607.

32. Barabási, A.-L. Network science. Philosophical Transactions of the Royal Society A: Mathematical, Physical and Engineering Sciences 2013, 371, 20120375.

33. Nguyen, A.-D. k-clique algorithm MATLAB Central File Exchange: MATLAB Central File Exchange, 2024.

34. Abdi, H.; Williams, L.J. Principal component analysis. Wiley interdisciplinary reviews: computational statistics 2010, 2, 433–459.

35. Kaiser, H.F. Computer program for varimax rotation in factor analysis. Educational and psychological measurement 1959, 19, 413–420.

36. Brier, G.W. Verification of forecasts expressed in terms of probability. Monthly weather review 1950, 78, 1–3.

37. Pencina, M.J.; D’Agostino Sr, R.B.; D’Agostino Jr, R.B.; Vasan, R.S. Evaluating the added predictive ability of a new marker: from area under the ROC curve to reclassification and beyond. Statistics in medicine 2008, 27, 157–172.

38. Derényi, I.; Palla, G.; Vicsek, T. Clique Percolation in Random Networks. Physical Review Letters 2005, 94, 160202, doi:10.1103/PhysRevLett.94.160202.

39. Porter, M.A.; Onnela, J.P.; Mucha, P.J. Communities in networks. Notices of the American Mathematical Society 2009, 56, 1082–1097.

40. Fortunato, S. Community detection in graphs. Physics Reports 2010, 486, 75–174, 10.1016/j.physrep.2009.11.002.

41. Katopodis, P.; Pappas, E.M.; Katopodis, K.P. Acid‒base abnormalities and liver dysfunction. Annals of Hepatology 2022, 27, 100675, 10.1016/j.aohep.2022.100675.

42. Walsh, T.S.; Mc Lellan, S.; Mackenzie, S.J.; Lee, A. Hyperlactatemia and Pulmonary Lactate Production in Patients With Fulminant Hepatic Failure. Chest 1999, 116, 471–476, 10.1378/chest.116.2.471.

43. Yudkin, J.; Cohen, R.D. The contribution of the kidney to the removal of a lactic acid load under normal and acidotic conditions in the conscious rat. Clinical science and molecular medicine 1975, 48, 121–131.

44. Bellomo, R. Bench-to-bedside review: lactate and the kidney. Crit Care 2002, 6, 322–326, doi:10.1186/cc1518.

45. Bihari, D.; Gimson, A.E.; Lindridge, J.; Williams, R. Lactic acidosis in fulminant hepatic failure. Some aspects of pathogenesis and prognosis. J Hepatol 1985, 1, 405–416, doi:10.1016/s0168-8278(85)80778-9.

46. Bernal, W.; Donaldson, N.; Wyncoll, D.; Wendon, J. Blood lactate as an early predictor of outcome in paracetamol-induced acute liver failure: a cohort study. Lancet 2002, 359, 558–563, doi:10.1016/s0140-6736(02)07743-7.

47. Riordan, S.M.; Williams, R. Blood lactate and outcome of paracetamol-induced acute liver failure. Lancet 2002, 360, 573; author reply 573-574, doi:10.1016/s0140-6736(02)09729-5.

48. Bernal, W. Lactate is important in determining prognosis in acute liver failure. Journal of Hepatology 2010, 53, 209–210, doi:10.1016/j.jhep.2010.02.017.

49. Schmidt, L.E.; Larsen, F.S. Blood lactate as a prognostic marker in acetaminophen-induced acute liver failure. Hepatology 2003, 37, 1199–1201, doi:10.1002/hep.510370530.

50. Alcorn, J. Arterial blood lactate measurements quickly identified risk for death from paracetamol-induced liver failure. ACP J Club 2002, 137, 117.

51. Scheiner, B.; Lindner, G.; Reiberger, T.; Schneeweiss, B.; Trauner, M.; Zauner, C.; Funk, G.-C. Acid‒base disorders in liver disease. Journal of Hepatology 2017, 67, 1062–1073, doi:10.1016/j.jhep.2017.06.023.

52. Bihari, D.; Gimson, A.E.S.; Lindridge, J.; Williams, R. Lactic acidosis in fulminant hepatic failure: Some aspects of pathogenesis and prognosis. Journal of Hepatology 1985, 1, 405–416, 10.1016/S0168-8278(85)80778-9.

53. Mikael, L.; Claire, V.; Òscar, M.; Juan González del, C.; Aitor, A.-A.; Javier, J.; Pascual, P.; Bruno, M. Are there differences in the relationship between respiratory rate and oxygen saturation between patients with COVID-19 and those without COVID-19? Insights from a cohort-based correlational study. Emergency Medicine Journal 2023, 40, 805, doi:10.1136/emermed-2022-212882.

54. Mower, W.R.; Sachs, C.; Nicklin, E.L.; Safa, P.; Baraff, L.J. A comparison of pulse oximetry and respiratory rate in patient screening. Respiratory Medicine 1996, 90, 593–599, 10.1016/S0954-6111(96)90017-7.

55. Oyelade, T.; Canciani, G.; Bottaro, M.; Zaccaria, M.; Formentin, C.; Moore, K.; Montagnese, S.; Mani, A.R. Heart rate turbulence predicts survival independently from severity of liver dysfunction in patients with cirrhosis. Frontiers in physiology 2020, 11, 602456.

56. Tan, Y.Y.; Montagnese, S.; Mani, A.R. Organ system network disruption is associated with poor prognosis in patients with chronic liver failure. Frontiers in physiology 2020, 11, 983.

57. D’Amico, G.; Garcia-Tsao, G.; Pagliaro, L. Natural history and prognostic indicators of survival in cirrhosis: a systematic review of 118 studies. Journal of hepatology 2006, 44, 217–231.

58. Asrani, S.K.; Kamath, P.S. Natural history of cirrhosis. Current gastroenterology reports 2013, 15, 1–6.

59. Geiger, B.C.; Kubin, G. Relative information loss in the PCA. In Proceedings of the 2012 IEEE Information Theory Workshop, 2012; pp. 562–566.

60. National Institute of, D.; Digestive; Kidney, D. LiverTox: clinical and research information on drug-induced liver injury; National Institute of Diabetes and Digestive and Kidney Diseases: 2012.

61. Hinson, J.A.; Roberts, D.W.; James, L.P. Mechanisms of acetaminophen-induced liver necrosis. Handb Exp Pharmacol 2010, 369–405, doi:10.1007/978-3-642-00663-0_12.

62. Yang, R.; Li, K.; Zou, C.; Wee, A.; Liu, J.; Liu, L.; Li, M.; Wu, T.; Wang, Y.; Ma, Z.;, et al. Alanine Aminotransferase and Bilirubin Dynamic Evolution Pattern as a Novel Model for the Prediction of Acute Liver Failure in Drug-Induced Liver Injury. Frontiers in Pharmacology 2022, 13.

63. Zimmerman, H.J. The spectrum of hepatotoxicity. Perspectives in biology and medicine 1968, 12, 135–161.

64. Hayashi, P.H.; Rockey, D.C.; Fontana, R.J.; Tillmann, H.L.; parenclitic owitz, N.; Barnhart, H.X.; Gu, J.; Chalasani, N.P.; Reddy, K.R.; Sherker, A.H. Death and liver transplantation within 2 years of onset of drug-induced liver injury. Hepatology 2017, 66, 1275–1285.

65. Robles-Diaz, M.; Lucena, M.; Kaplowitz, N.; Stephens, C.; Medina-Cáliz, I.; González-Jimenez, A.; Ulzurrun, E.; Gonzalez, A.; Fernandez, M.; Romero-Gómez, M. Spanish DILI Registry; SLatinDILI Network; Safer and Faster Evidence-based Translation Consortium. Use of Hy’s law and a new composite algorithm to predict acute liver failure in patients with drug-induced liver injury. Gastroenterology 2014, 147, 109–118.

66. Garcia-Cortes, M.; Robles-Diaz, M.; Stephens, C.; Ortega-Alonso, A.; Lucena, M.I.; Andrade, R.J. Drug induced liver injury: an update. Archives of Toxicology 2020, 94, 3381–3407, doi:10.1007/s00204-020-02885-1.

67. Alves, P.S.; Camilo, E.A.; Correia, J.P. The SGOT/SGPT ratio in alcoholic liver disease. Acta Med Port 1981, 3, 255–260.

68. Nyblom, H.; Berggren, U.; Balldin, J.; Olsson, R. HIGH AST/ALT RATIO MAY INDICATE ADVANCED ALCOHOLIC LIVER DISEASE RATHER THAN HEAVY DRINKING. Alcohol and Alcoholism 2004, 39, 336–339, doi:10.1093/alcalc/agh074.

69. Nyblom, H.; Björnsson, E.; Simrén, M.; Aldenborg, F.; Almer, S.; Olsson, R. The AST/ALT ratio as an indicator of cirrhosis in patients with PBC. Liver International 2006, 26, 840–845, 10.1111/j.1478-3231.2006.01304.x.

70. Larsen, F.S.; Hansen, B.A.; Blei, A.T. Intensive care management of patients with acute liver failure with emphasis on systemic hemodynamic instability and cerebral edema: a critical appraisal of pathophysiology. Can J Gastroenterol 2000, 14 Suppl D, 105d–111d, doi:10.1155/2000/493629.

71. Trewby, P.; Williams, R. Pathophysiology of hypotension in patients with fulminant hepatic failure. Gut 1977, 18, 1021–1026.

72. Ellis, A.; Wendon, J. Circulatory, respiratory, cerebral, and renal derangements in acute liver failure: pathophysiology and management. In Proceedings of the Seminars in liver disease, 1996; pp. 379–388.

73. Ring-Larsen, H.; Palazzo, U. Renal failure in fulminant hepatic failure and terminal cirrhosis: a comparison between incidence, types, and prognosis. Gut 1981, 22, 585–591, doi:10.1136/gut.22.7.585.

74. Wilkinson, S.; Blendis, L.; Williams, R. Frequency and type of renal and electrolyte disorders in fulminant hepatic failure. Br Med J 1974, 1, 186–189.

75. Moore, K. Renal failure in acute liver failure. Eur J Gastroenterol Hepatol 1999, 11, 967–975, doi:10.1097/00042737-199909000-00004.

76. Siniscalchi, A.; Dante, A.; Spedicato, S.; Riganello, L.; Zanoni, A.; Cimatti, M.; Pierucci, E.; Bernardi, E.; Miklosova, Z.; Moretti, C.; Faenza, S. Hyperdynamic circulation in acute liver failure: reperfusion syndrome and outcome following liver transplantation. Transplant Proc 2010, 42, 1197–1199, doi:10.1016/j.transproceed.2010.03.097.

77. Oldenbeuving, G.; McDonald, J.R.; Goodwin, M.L.; Sayilir, R.; Reijngoud, D.J.; Gladden, L.B.; Nijsten, M.W. A patient with acute liver failure and extreme hypoglycaemia with lactic acidosis who was not in a coma: causes and consequences of lactate-protected hypoglycaemia. Anaesth Intensive Care 2014, 42, 507–511, doi:10.1177/0310057x1404200413.

78. Record, C.O.; Iles, R.A.; Cohen, R.D.; Williams, R. Acid‒base and metabolic disturbances in fulminant hepatic failure. Gut 1975, 16, 144–149, doi:10.1136/gut.16.2.144.

79. Talari, K.; Goyal, M. Retrospective studies–utility and caveats. Journal of the Royal College of Physicians of Edinburgh 2020, 50, 398–402.

80. Moss, T.J.; Lake, D.E.; Calland, J.F.; Enfield, K.B.; Delos, J.B.; Fairchild, K.D.; Moorman, J.R. Signatures of Subacute Potentially Catastrophic Illness in the ICU: Model Development and Validation. Crit Care Med 2016, 44, 1639–1648, doi:10.1097/ccm.0000000000001738.

81. Gheorghita, M.; Wikner, M.; Cawthorn, A.; Oyelade, T.; Nemeth, K.; Rockenschaub, P.; Gonzalez Hernandez, F.; Swanepoel, N.; Lilaonitkul, W.; Mani, A.R. Reduced oxygen saturation entropy is associated with poor prognosis in critically ill patients with sepsis. Physiological Reports 2022, 10, e15546.

