## Supplementary material for "Application of physiological network mapping in the prediction of survival in critically ill patients with acute liver failure": Table S 1

### Supplementary Materials

***Table S 1.*** *Principal components after Varimax and rotation and Kaiser Normalization. The KMO (Kaiser-Meyer-Olkin) test that the sample is adequate for PCA showed p values < 0.001 (Chi-Square = 1844.299, p-value = <0.001).*

| | **Principal Components** | | | | | | | | |
|---|---|---|---|---|---|---|---|---|---|
| **Variables** | 1 | 2 | 3 | 4 | 5 | 6 | 7 | 8 | 9 |
| Alanine Aminotransferase | 0.828 | | | | | | | | |
| Aspartate Transaminase | 0.817 | | | | | | | | |
| International Normalized Ratio | 0.764 | | | | | | | | |
| Oxygen Saturation | | | | | | | | | |
| Serum Creatinine | | 0.845 | | | | | | | |
| Urea | | 0.807 | | | | | | | |
| Phosphate | | 0.69 | | | | | | | |
| Serum Albumin | | | 0.692 | | | | | | |
| Mean Blood Pressure | | | 0.649 | | | | | | |
| Haemoglobin | | | 0.607 | | | | | | |
| Alkaline Phosphatase | | | | | | | | | |
| Serum Sodium | | | | 0.924 | | | | | |
| Chloride | | | | 0.831 | | | | | |
| Bicarbonate | | | | | 0.801 | | | | |
| Blood pH | | | | | 0.765 | | | | |
| Heart Rate | | | | | | 0.815 | | | |
| Respiratory Rate | | | | | | 0.707 | | | |
| Blood Glucose | | | | | | | 0.809 | | |
| Lactate | | | | | | | 0.571 | | |
| Glasgow Coma Score | | | | | | | | | |
| Platelet Count | | | | | | | | 0.836 | |
| White Blood Count | | | | | | | | 0.757 | |
| Total Bilirubin | | | | | | | | | 0.744 |
| Body Temperature | | | | | | | | | -0.728 |

*Na; Serum Sodium, Cl; chloride, AST; aspartate transaminase, ALT; alanine aminotransferase, GCS; Glasgow Coma Score, Bil; Total Bilirubin, ALP; Alkaline Phosphatase, Cr; Serum Creatinine, Na; Serum Sodium, Glu; Blood Glucose, HR; Heart Rate, Temp; Temperature, SEM; Standard Error of Mean, HR; Hazard Ratio, CI; Confident Interval.*
