## Supplementary material for "Application of physiological network mapping in the prediction of survival in critically ill patients with acute liver failure": Table S 2

### Supplementary Materials

***Table S 1.*** *Univariate Cox regression analysis of the Principal Components based on ICU survival of patients.*

| Principal component | β | SEM | Hazard Ratio (95% CI) | p-value |
|---|---|---|---|---|
| PC-1 | 0.255 | 0.092 | 1.08 (1.29 - 1.55) | **0.005** |
| PC-2 | 0.168 | 0.097 | 0.98 (1.18 - 1.43) | 0.082 |
| PC-3 | -0.258 | 0.093 | 0.64 (0.77 - 0.93) | **0.005** |
| PC-4 | 0.13 | 0.106 | 0.93 (1.14 - 1.4) | 0.222 |
| PC-5 | -0.084 | 0.102 | 0.75 (0.92 - 1.12) | 0.414 |
| PC-6 | 0.111 | 0.107 | 0.91 (1.12 - 1.38) | 0.299 |
| PC-7 | 0.226 | 0.103 | 1.03 (1.25 - 1.53) | **0.027** |
| PC-8 | -0.070 | 0.099 | 0.77 (0.93 - 1.13) | 0.478 |
| PC-9 | -0.070 | 0.099 | 0.77 (0.93 - 1.13) | 0.478 |

*SEM; Standard Error of Mean, CI; Confident Interval.*
